# When Campus Feeds the Clock: How the University Food Environment Shapes the Circadian Rhythms of Undergraduate Students at the University of Medical Sciences, Ondo

**DOI:** 10.64898/2026.09.20.26363532

**Authors:** Oluwabunmi Emmanuel Folorunso, Joy Chinaza Amadi, Oluwaseun Funmi Akinmoladun, Beatrice Olubukola Ogunba

## Abstract

**Background:** Circadian eating patterns, referring to the timing and regularity of meal intake relative to the body’s internal clock, are increasingly recognized as important determinants of metabolic health among university students. However, empirical evidence on how the university food environment influences circadian eating patterns remains scarce, particularly within Nigerian tertiary institutions. This study assessed the influence of the university food environment on the circadian eating patterns of undergraduate students at the University of Medical Sciences, Ondo (UNIMED).

**Methods:** A descriptive cross-sectional study was conducted among 302 undergraduate students selected through a multistage sampling technique. Data were collected using a structured, facilitator-administered questionnaire assessing socio-demographic characteristics, the university food environment (UNI-FOOD, across six domains), circadian eating patterns, influencing factors, and anthropometric measurements. Data were analysed using SPSS version 27, employing descriptive statistics, chi-square tests, and univariable and multivariable binary logistic regression, with significance set at p < 0.05.

**Results:** The mean age of respondents was 20.03 ± 2.58 years, and the majority were female (68.9%). Students generally perceived the university food environment as moderate, with affordability rated the weakest domain (mean 2.65 ± 0.51) and food promotion/visibility the strongest (3.27 ± 0.52). Most respondents (71.9%) exhibited moderate circadian eating patterns, while 21.2% had poor patterns. A significant association was found between the overall food environment and circadian eating patterns (χ² = 44.369, p < 0.001). After adjustment, poor physical food accessibility (AOR = 6.44, 95% CI: 2.00–20.72, p = 0.002) and a poor food policy environment (AOR = 9.53, 95% CI: 3.51–25.87, p < 0.001) were independent predictors of poor circadian eating patterns. Circadian eating pattern was also significantly associated with BMI category (χ² = 7.346, p = 0.025).

**Conclusion:** The university food environment, particularly physical accessibility and food policy, significantly influences circadian eating patterns among undergraduate students. Improving campus food accessibility and implementing supportive food policies may promote healthier meal timing and nutritional outcomes.

## Background of the study

Circadian eating patterns involve timing your meals to match your body’s internal 24-hour clock to improve digestion, metabolism, and overall health. It focuses on when people eat, not just what was eaten (1). The principle involves consuming most calories during daylight hours and limiting late-night eating (2). This approach is grounded in the body’s circadian rhythm, which regulates metabolism, insulin sensitivity, and hormone production. The human body is governed by endogenous circadian rhythms, which is approximately 24-hour cycles that regulate various physiological processes, including hormone secretion, glucose metabolism, and energy expenditure. These rhythms are orchestrated by a central pacemaker in the suprachiasmatic nucleus (SCN) of the hypothalamus and are influenced by external cues such as light exposure and feeding cycles (Alum, 2025). When feeding behavior is misaligned with these circadian signals, as observed in shift workers, individuals with irregular eating schedules, or those who engage in late-night eating, there is a heightened risk of weight gain, insulin resistance, and cardiometabolic dysfunction (4). While caloric content and macronutrient composition remain foundational, accumulating research demonstrates that when we eat is equally critical. Time-restricted eating (TRE), early time-restricted feeding (eTRE), and other temporal strategies have shown promising effects on glycemic control, blood pressure, inflammatory markers, and body weight even in the absence of calorie reduction (5). This emerging paradigm challenges the long-standing notion that “a calorie is a calorie,” highlighting the need to integrate temporal patterns into dietary guidelines and interventions (6). Unlike traditional dietary interventions that focus solely on the quantity and quality of food, circadian nutrition introduces the dimension of temporal alignment, which may provide a low-cost, behaviorally feasible, and metabolically useful approach to noncommunicable disease prevention and treatment among the general population, most especially among university students (5).

The university years represent a critical period in the transition from adolescence to adulthood and for the development of long-term eating habits, which can influence both short-term and chronic health outcomes (7). University students represent a population particularly susceptible to circadian eating disruption, as academic demands, irregular class schedules, and the transition to independent living often lead to skipped meals, delayed breakfast, and frequent late-night eating (7). These patterns misalign food intake with the body’s internal clock, a state increasingly linked to impaired glucose regulation and metabolic dysfunction independent of overall caloric intake (8). Given the rising burden of non-communicable diseases among young adults, understanding circadian eating patterns in this population offers an important, underexplored avenue for early risk identification and intervention. Recent evidence highlights the growing trend of consuming the majority of daily calories later in the day; this behavior was associated with a higher risk of metabolic disorders. Individuals with an evening chronotype are approximately 2.5 times more likely to develop type 2 diabetes compared to morning chronotypes, even after adjusting for sleep duration. Moreover, evening chronotypes tend to have delayed sleep patterns, which may further impact metabolic regulation (9). A significant proportion of university students engage in unhealthy eating patterns at night. Previous research indicates that 57.10% of university students regularly consume late-night snacks, with 6.79% eating late-night snacks every night, 23.15% eating late-night snacks frequently, and 27.16% eating late-night snacks occasionally (7).

In Nigeria, this phenomenon is increasingly driven by intense traffic gridlocks (forcing late-night dinners), demanding shift work (among healthcare, factory, and security personnel), and a rapidly growing 24/7 economy. Poor circadian eating patterns including skipped breakfasts, late-night eating, and erratic meal timing, disrupt the body’s internal biological clock and are associated with significant academic, metabolic, and psychological consequences among Nigerian university students (10). Irregular eating lowers blood glucose and impairs concentration and memory retention, while late-night eating disrupts sleep quality, contributing to daytime fatigue and reduced classroom performance (11). These disruptions are compounded by poor dietary choices often cheap, energy-dense, and nutritionally imbalanced that place students at risk of a double burden of malnutrition and gastrointestinal complaints such as acid reflux and ulcers (12). Psychologically, irregular eating and poor sleep synergistically heighten stress reactivity and worsen depressive and anxiety symptoms, particularly during periods of academic pressure (10).

Several studies suggest that prioritizing morning energy intake while reducing evening calorie consumption may protect against metabolic syndrome (MetS), even in individuals with normal body weight (9)

Early time-restricted eating, involving finishing dinner by 6 to 7 pm, has been shown to improve insulin sensitivity, fasting glucose, blood pressure, and lipid profile (5). Meta-analyses and clinical trials show that time-restricted eating leads to reductions in body weight, fat mass, and waist circumference, even without strict calorie restriction (5). Previous studies also indicate that aligning eating patterns with circadian rhythms may reduce the risk of obesity, diabetes, cardiovascular disease and age-related decline (Flanagan et al., 2021, Regmi & Heilbronn, 2020) as avoiding late-night meals can also enhance sleep quality and maintain hormonal balance (6).

To achieve the focus and memory necessary to attain academic success, students need well-planned diets that are balanced in energy, essential nutrients and timely. However, university life has often been associated with poor eating habits in students due to academic demands and a poor food environment (15). Poor food selection is particularly a problem among students, as they often prioritise sensory appeal, price, convenience, and availability over the healthfulness of their diets (Ekerette & Udo, 2025). The university food environment is generally characterized by limited access to fresh, nutrient-dense meals, with availability skewed toward inexpensive, convenient, and highly processed options; campus canteens, vending machines, and surrounding food vendors typically prioritize affordability and speed over nutritional quality, offering extended or round-the-clock access to fast food, sugary beverages, and energy-dense snacks (16). This environment, shaped by the practical constraints of student life limited budgets, packed academic schedules, and irregular daily routines creates conditions that make convenient, poorly timed, and nutritionally inadequate eating the default rather than the exception (17).

These structural conditions are closely linked to poor circadian eating patterns among students (7). Irregular academic schedules, late-night studying, and round-the-clock access to fast food and ultra-processed snacks over fresh produce push students toward opportunistic, poorly timed eating rather than consistent, circadian-aligned meal patterns, while the absence of structured, affordable meal options in the early morning contributes to widespread breakfast skipping; social and academic pressures further encourage eating outside natural daylight hours (18). This environmentally driven irregularity disrupts chrononutrition, as meal timing—alongside light exposure serves as a key synchronizer of peripheral metabolic clocks in organs such as the liver and gut (19). Eating late at night contradicts the body’s natural nocturnal decline in metabolism, reducing insulin sensitivity and impairing glucose tolerance, while also suppressing melatonin release and blunting satiety signals from leptin, elevating hunger-driving ghrelin, and prolonging cortisol activity collectively heightening obesity and metabolic risk (20). Research suggests that meal timing can significantly influence metabolic health. Consuming meals during the body’s active phase, typically earlier in the day, aligns with peak insulin sensitivity and glucose tolerance (21). Conversely, late-night eating has been associated with impaired glucose metabolism and increased fat storage (22). These findings highlight the potential of chrononutrition as a complementary approach to traditional dietary strategies. Together, these findings position the university food environment as a critical structural driver of circadian eating disruption and associated metabolic vulnerability among students(23).

Despite growing global evidence linking circadian eating disruption to metabolic, cognitive, and psychological outcomes (24), most existing studies have been conducted in Western or Asian university settings, with limited data from Nigerian tertiary institutions. Furthermore, while previous Nigerian studies have examined dietary patterns or food insecurity among students in isolation (Iheme et al., 2026,Sholeye OO et al., 2021) few have specifically investigated how the structural characteristics of the university food environment such as canteen hours, food vendor accessibility, and affordability of healthy options directly shape students’ meal timing and circadian eating behavior. This gap is particularly pronounced among students in health-focused institutions such as the University of Medical Sciences, Ondo, where students face demanding academic and clinical schedules yet paradoxically possess greater theoretical knowledge of nutrition and health, raising unanswered questions about whether such knowledge translates into circadian-aligned eating practices when environmental and structural constraints are considered (13). Studies empirically examining the influence of the university food environment on circadian eating patterns among undergraduates within this specific context remain scarce; hence, this study aims to assess the influence of the university food environment on the circadian eating patterns of undergraduate students of the University of Medical Sciences, Ondo.

## Methodology

### Study Design and Settings

The design for the research was descriptive cross-sectional, conducted in Ondo town, ondo state, Nigeria. Ondo State is situated in the southwestern part of the country, specifically in Ondo State’s southern region. Ondo, Nigeria. The latitude and longitude coordinates are 7.100005, 4.841694. The city is approximately 54 kilometres (34 miles) northeast of Akure, the capital city of Ondo State. The state lies between longitudes 4″″ 30″″ and 6″″ East of the Greenwich Meridian 5″ ″ 45″″ and 8″″ 15″″ North of the Equator. Population 3,460,877 comprising 1,745,057 Males and 1,715,820 Females (as at 2006) and projected population of 4,883,792 comprising 2,462,525 males and, 2,421,261 females (by the State Bureau of Statistics). Ondo Town, the study setting, is the second-largest city in Ondo State, Nigeria, and serves as a major commercial and agricultural hub, particularly for cocoa production. Situated in southwestern Nigeria near the southern edge of the Yoruba Hills, the town is a notable trade center for cocoa, yams, cassava, and palm oil, and is also known for its traditional Aso Oke fabric weaving. Ondo Town hosts several higher education institutions, including the University of Medical Sciences, Ondo, and Adeyemi Federal University of Education. The town retains strong traditional heritage, being ruled by the Osemawe of Ondo, the paramount monarch whose lineage traces back to the historic Ondo Kingdom, and features prominent landmarks such as the Ondo Central Market (Oja Oba), located near the palace.

The University of Medical Sciences (UNIMED), Ondo, is a specialized medical and health sciences university located along Laje Road, Ondo City, Ondo State, Nigeria. Established in 2015, it is Nigeria’s first specialized medical university and provides undergraduate and postgraduate education in medical, health, and related scientific disciplines. The university comprises several academic units, including the Faculties of Clinical Sciences, Dental Sciences, Nursing Sciences, Allied Health Sciences, Basic Medical Sciences, Basic Clinical Sciences, Medical Rehabilitation, and Sciences, as well as the School of Public Health. These faculties encompass numerous departments and academic programmes covering medicine, dentistry, nursing, allied health professions, Basic medical sciences, rehabilitation sciences, and Sciences. The university has a student population of over 8,000 currently enrolled students, with total enrolment exceeding 9,000 students.

### Study Population

The study population comprised male and female undergraduate students aged 18–30 years.

### Sample Size Determination

The sample size for this study was determined using Le Fisher’s formula for estimating sample size in descriptive cross-sectional studies (Fisher et al., 1998). A prevalence of 11.6% obesity among Nigerian adolescents and young adults (Adeloye et al., 2021) was used.

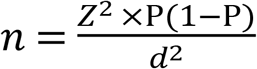

Where n = the desired sample size

Z = The standard normal deviate corresponding to the desired confidence level (1.96 for a 95% confidence level)

P = estimated prevalence of obesity (0.116)

1 − P = complement of the estimated prevalence (0.884)

d = desired margin of error (0.038)

#### 3.4.1 Sample Size Calculation

Using:

P = 0.116

95% confidence level (Z = 1.96)

Margin of error (d = 0.038)

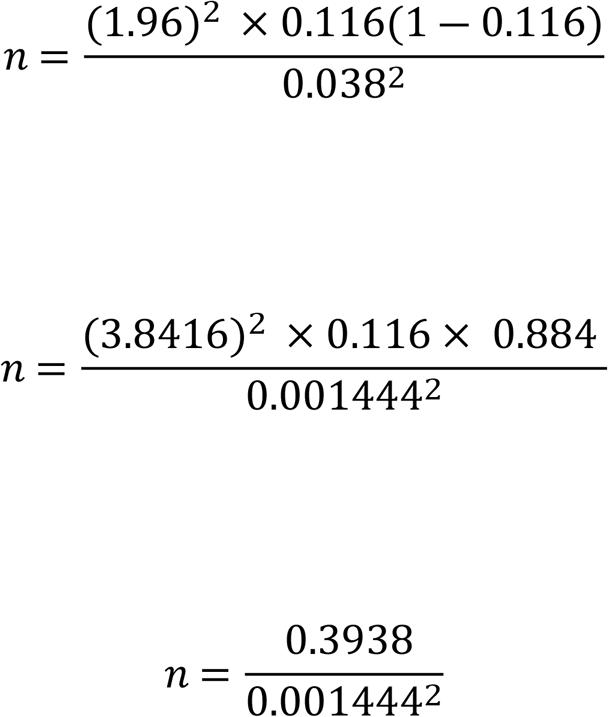

n = 272.71 ≈ 273

Therefore, the minimum required sample size was 273 respondents.

**Note:** To guarantee there would be enough final data for analysis, the computed sample size was increased by an expected non-response or dropout rate of 10%.

10% x 273 = 27.3

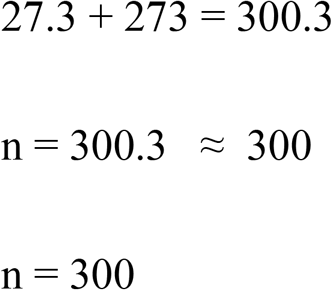

### Sample Size Procedure

A multistage technique was adopted to select respondents for this study. Using this sampling technique required step by step considerations. In stage one, Ondo town consist of two local government which are the Ondo east (Bolorunduro) and Ondo west (Ondo city). Ondo west was purposively selected for this research because it consists of the main tertiary institutions in Ondo town. Stage two; Ondo West consists of the main tertiary institutions which include: University of Medical Sciences, Ondo, Adeyemi Federal University of Education, and Wesley University. The University of Medical Sciences, Ondo was randomly selected for this study. Stage three; The university consists of eight faculties offering various undergraduate programmes from which respondents for the study were selected, the faculties namely the Faculty of Basic Medical Sciences (Anatomy, Physiology and Biochemistry), the Faculty of Clinical Sciences (Medicine and Surgery), the Faculty of Basic Clinical Sciences (Anatomic Pathology, Chemical Pathology, Medical Microbiology and Pharmacology), the Faculty of Dental Sciences (Dentistry, Dental Therapy and Dental Technology), the Faculty of Nursing Science (Nursing Science), the Faculty of Science (Biology, Chemistry, Physics and Mathematics), the Faculty of Allied Health Sciences (Medical Laboratory Science, Human Nutrition and Dietetics, Radiography, Health Information Management, and Complementary and Alternative Medicine), the Faculty of Medical Rehabilitation (Physiotherapy, Prosthetics and Orthotics, Audiology and Occupational Therapy), and the Faculty of Pharmacy (Pharmacy).

From the faculties mentioned above, three faculties were selected, using simple random sampling. Thereafter, two departments were selected from each of the selected faculties using simple random sampling, giving a total of six departments. The selected faculties were the Faculty of Clinical Sciences, Faculty of Allied Health Sciences, and Faculty of Medical Rehabilitation. The selected departments included (Medicine and Surgery [MBBS]) and (Dentistry) from the Faculty of Clinical Sciences; (Medical Laboratory Science) and (Radiography and Radiation Science) from the Faculty of Allied Health Sciences; and (Physiotherapy) and (Prosthetics and Orthotics) from the Faculty of Medical Rehabilitation

Stage four; the total of 50 students were selected from each of the six departments to arrive at a total sample size of 300 respondents. Respondents were then selected from each departments using simple random sampling. Where the selected departments had approximately equal students populations. This approach ensured that the sample was representative of the undergraduate student population and that every eligible students had equal chance of being selected

### Data Collection Instrument

The instrument used for data collection was a structured, facilitated-administered questionnaire designed to obtain relevant information from respondents. The questionnaire was divided into five (5) sections:

Section A collected socio-demographic characteristics such as age, gender, level of study, residence, monthly food allowance, and parental occupation. Section B assessed the university food environment across key domains, including food availability, temporal (circadian) accessibility, affordability, physical accessibility, promotion and visibility, and the policy environment. Section C assessed circadian eating patterns, including meal timing, frequency, and consistency of eating behaviour. Section D examined factors influencing eating patterns, including academic stress, financial constraints, and campus living conditions. Section E collected anthropometric measurements, including weight, height, and waist circumference, for the assessment of nutritional status.

### Measurement of variables

This study assessed key variables related to the influence of the university food environment on the circadian eating patterns of undergraduate students at the University of Medical Sciences, Ondo. Data were collected using a structured, facilitated questionnaire designed specifically to address the study objectives. The variables measured included socio-demographic characteristics, the university food environment, circadian eating patterns, factors influencing eating patterns, and anthropometric measurements.

The university food environment was assessed using (UNI-FOOD) was assessed across six domains: food availability, temporal accessibility, affordability, physical accessibility, promotion and visibility, and the policy environment (26). Responses were measured using a five-point Likert scale: Strongly Agree (5), Agree (4), Neutral (3), Disagree (2), and Strongly Disagree (1). Negatively worded statements were reverse-scored to ensure consistency in interpretation. Scores for each domain were summed to generate composite scores, which were categorized into poor, moderate, and good food environments.

Circadian eating patterns were measured using indicators such as meal timing (breakfast, lunch, and dinner), meal frequency, late-night eating, meal regularity, and meal consistency. Higher scores indicated healthier and more regular circadian eating patterns.

Factors influencing eating patterns were assessed using questions on academic stress, financial constraints, and campus living conditions. Responses were scored using the same five-point Likert scale, with higher scores indicating a greater influence of these factors on students’ eating patterns.

Anthropometric measurements were obtained using respondents’ weight (kg), height (m), and waist circumference (cm). Body Mass Index (BMI) was calculated using the formula:

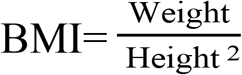

BMI was classified according to the World Health Organization (WHO) classification as underweight (<18.5 kg/m²), normal weight (18.5–24.9 kg/m²), overweight (25.0–29.9 kg/m²), and obese (≥30.0 kg/m²).

### Pretesting, Validity, and Reliability of the Research Instrument

Before the main data collection, the questionnaire was pretested among undergraduate students at the Osun state University, Osogbo, Osun State, Nigeria, an institution outside the study area. A pretest sample representing 10% of the total sample size was utilised. The purpose of this exercise was to identify and resolve any unclear or ambiguous items and to confirm that the questions were suitable and appropriate for the intended study population

Also, the university food environment was assessed using the University Food Environment Questionnaire (UNI-FOOD), an adopted instrument that evaluates six domains: food availability, temporal (time) accessibility, affordability, physical accessibility, promotion and visibility, and the policy environment. In addition to UNI-FOOD, items assessing institutional factors and circadian eating patterns were researcher-constructed specifically for the purpose of this study. All subscales, whether adopted or researcher-constructed, were subjected to a reliability test using Cronbach’s alpha to establish internal consistency prior to administration, yielding coefficients ranging from acceptable to excellent: Food Availability (α = 0.716), Time Availability (α = 0.706), Affordability (α = 0.698), Physical Accessibility (α = 0.707), Food Promotion (α = 0.588), Food Policy Environment (α = 0.900), Circadian Eating Pattern (α = 0.614), and Institutional Factors (α = 0.719).

**Table 1:** Reliability Test Using Cronbach’s Alpha.

| Scale | Cronbach's $\alpha$ | Status |
| --- | --- | --- |
| Food Availability (FA) | 0.716 | ✓ Complete |
| Time Availability (TA) | 0.706 | ✓ Complete |
| Affordability (AF) | 0.698 | ✓ Complete |
| Physical Accessibility (PA) | 0.707 | ✓ Complete |
| Food Promotion (PV) | 0.588 | ✓ Complete |
| Food Policy Environment (PE) | 0.900 | ✓ Complete |
| Circadian Eating Pattern (CEP) | 0.614 | ✓ Complete |
| Institutional Factors (IF) | **0.719** | ✓ Complete |

### Inclusion Criteria

Participants were currently registered full-time undergraduate students of the University of Medical Sciences (UNIMED), Ondo, at the time of the study. They have completed at least one academic semester in the institution to ensure adequate exposure to the campus food environment and its influence on eating behaviour. In addition, only students physically present in Ondo City during the data collection period were included to ensure their eating patterns reflect the local university food environment.

### Exclusion Criteria

Part-time, sandwich, and postgraduate students were excluded from the study due to differences in academic schedules and duration of exposure to the university food environment compared to full-time undergraduates. Students who are absent from campus due to industrial training, rural clinical postings, or other external assignments outside Ondo City were also excluded, as they are not currently exposed to the study environment. In addition, students with diagnosed eating disorders, chronic illnesses that affect meal timing (such as Type 1 Diabetes), or physical disabilities that may interfere with accurate anthropometric measurements were excluded to ensure data validity and homogeneity.

### Data Analysis plan

Data collected were entered, cleaned, and analysed using the Statistical Package for the Social Sciences (SPSS) version 27. Data cleaning involved checking for missing values, inconsistencies, and outliers prior to analysis to ensure data quality and accuracy. Descriptive statistics, including frequencies, percentages, means, and standard deviations, were used to summarise respondents’ sociodemographic characteristics and key study variables, including university food environment domains and circadian eating patterns. Inferential statistics were employed to examine relationships between variables: chi-square tests were used to assess associations between categorical variables, while univariable and multivariable binary logistic regression analyses were conducted to determine the influence of the university food environment domains on circadian eating patterns among respondents. Variables significant at the univariable level were entered into the multivariable model to control for potential confounders and identify independent predictors, with results presented as crude and adjusted odds ratios (cOR/aOR) alongside 95% confidence intervals. Statistical significance for all analyses was set at p < 0.05.

### Ethical consideration

For this study, the participant recruitment period was 4th June 2026 – 30th June 2026. Ethical clearance for this study was granted by the Ethics Review Committee of the University of Medical Science, Ondo, Ondo State under reference number UNIMED-HREC/APV/2026/805. The study was carried out in full compliance with the ethical principles outlined in the Declaration of Helsinki and its subsequent amendments, as well as comparable ethical frameworks(39)

Prior to enrolment, all participants provided informed consent. Each participant was briefed on the purpose of the study and its potential benefits before completing and signing the consent form. Participants were explicitly informed of their right to voluntarily join or withdraw from the study at any point without any consequence. Among the benefits extended to participants was nutrition counselling on dietary pattern which served to raise awareness of the associated health implications.

Strict confidentiality was maintained throughout the study. All data were anonymised and secured on a password-protected device, accessible only to authorised members of the research team.

### Limitations of the Study

The Food Promotion subscale demonstrated a relatively low internal consistency (α = 0.588), falling slightly below the conventional threshold of 0.60–0.70 typically considered acceptable; this suggests the items within this subscale may not have consistently measured a single underlying construct, and findings related to food promotion should therefore be interpreted with some caution.

## RESULTS

### Socio-demographic characteristics of the respondents

This study investigated the influence of the university food environment on the circadian eating patterns of undergraduate students in Ondo west, Ondo Town. Table 2a revealed 302 undergraduate students with a mean age of 20.03 ± 2.58 years, ranging from 16 to 32 years participated in this study. More than half of the respondents 169(56.0%) were aged 20 years or older, while 133(44.0%) were younger than 20 years. Females made up the majority of the participants 208(68.9%), whereas males accounted for 94(31.1%). Most respondents were of Yoruba ethnicity 243(80.5%), majority identified as Christians 288(95.4%), while only 14(4.6%) practiced Islam. Regarding accommodation, over half of the respondents 161(53.3%) lived in off-campus apartments, 129(42.7%) resided in school hostels, and only 12(4.0%) lived with their parents.

**Table 2a.**
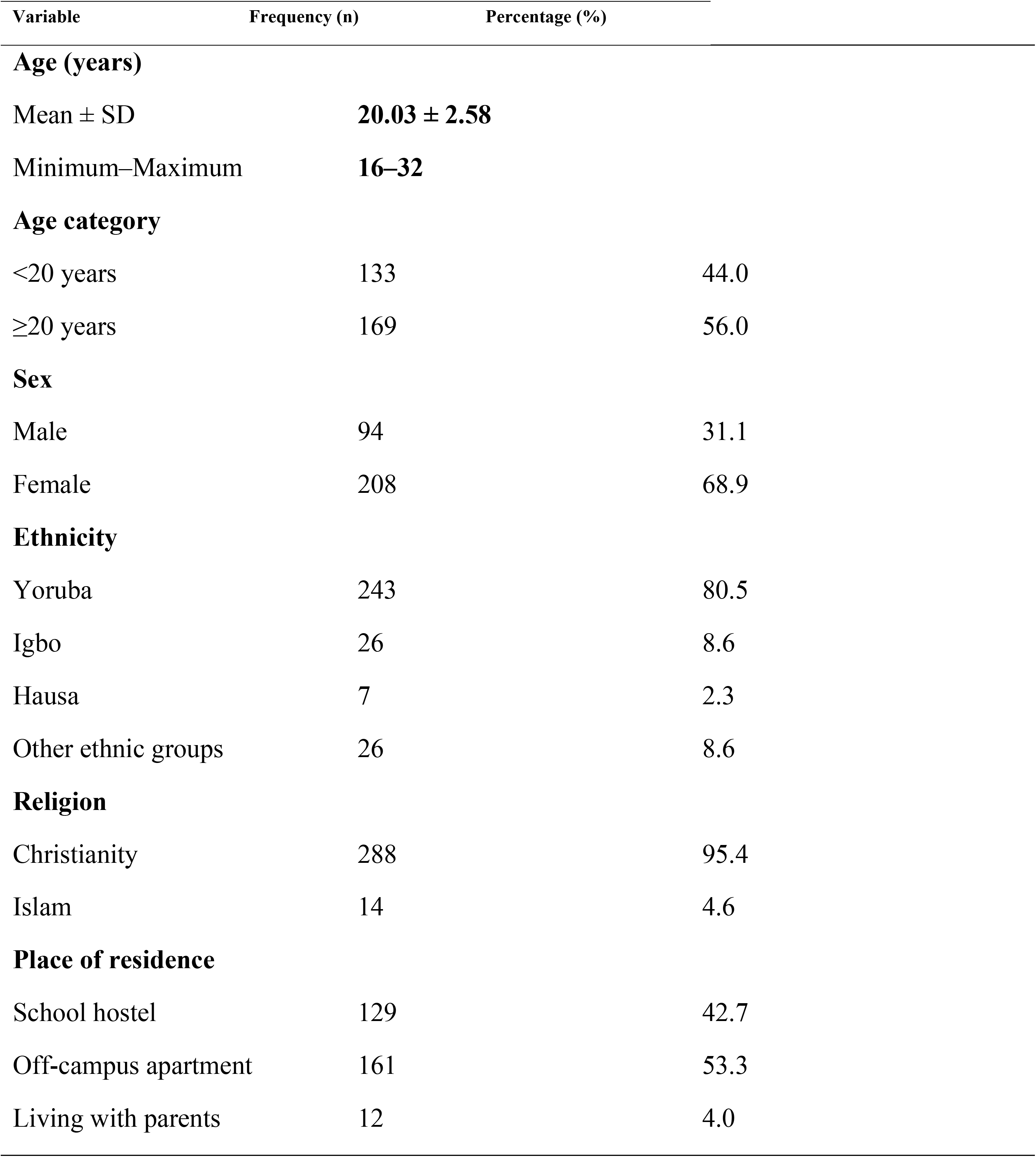
Socio-demographic characteristics of the respondents (N = 302)

### Socio-demographic characteristics of the respondents (2)

Table 2b indicated approximately 181(59.9%) reported receiving a monthly food allowance of ₦26,000–₦50,000, while only 18(6.0%) received ₦76,000 or more. Among the 300 respondents who provided information on living arrangements, 140(46.7%) lived with roommates, 110(36.7%) lived alone, and 50(16.7%) lived with their parents. Most respondents reported that both their mothers 231(76.5%) and fathers 253(83.8%) had attained tertiary education. Regarding parental occupation, 177(58.6%) of mothers were self-employed compared with 154(51.0%) of fathers who were government workers

**Table 2b.** Socio-demographic characteristics of the respondents (N = 302)

| Variable | Frequency (n) | Percentage (%) |
| --- | --- | --- |
| **Monthly food allowance (₦)** |  |  |
| ≤25,000 | 43 | 14.2 |
| 26,000–50,000 | 181 | 59.9 |
| 51,000–75,000 | 60 | 19.9 |
| ≥76,000 | 18 | 6.0 |
| **Living arrangement (n = 300)** |  |  |
| Alone | 110 | 36.7 |
| With parents | 50 | 16.7 |
| With roommate(s) | 140 | 46.7 |
| **Mother's educational level** |  |  |
| Primary/Secondary | 71 | 23.5 |
| Tertiary | 231 | 76.5 |
| **Father's educational level** |  |  |
| Primary/Secondary | 49 | 16.2 |
| Tertiary | 253 | 83.8 |
| **Mother's occupation** |  |  |
| Government worker | 125 | 41.4 |
| Self-employed | 177 | 58.6 |
| **Father's occupation** |  |  |
| Government worker | 154 | 51.0 |
| Self-employed | 148 | 49.0 |

### Mean scores of the university food environment domains among undergraduate students

The university food environment was assessed across six domains: food availability, timing, affordability, physical accessibility, food promotion/visibility, and food policy environment. Table 3 showed the domain mean scores ranged from 2.65 to 3.27 on a five-point scale, indicating moderate perceptions of the university food environment, with affordability (2.65), emerging as the weakest aspect and promotion/visibility as the strongest (3.27). As shown in Table 2, the Food Promotion/Visibility domain had the highest mean score (3.27 ± 0.52), indicating that students generally perceived food promotion and visibility practices on campus more favourably than the other aspects of the food environment. This was followed by the Food Policy Environment (2.99 ± 0.71) and Physical Accessibility (2.89 ± 0.50) domains. The Timing domain had a mean score of 2.82 ± 0.50, while Food Availability had a comparable mean score of 2.76 ± 0.52. The Affordability domain recorded the lowest mean score (2.65 ± 0.51), suggesting that the cost of healthy foods was perceived as the greatest challenge within the university food environment. Interpretation. While students generally acknowledged the visibility and promotion of foods within the university environment, they perceived the affordability of healthier food options as relatively poor. This finding is likely to be important when discussing the relationship between the food environment and students’ circadian eating patterns and nutritional status later in the manuscript.

**Table 3.** Mean scores of the university food environment domains among undergraduate students (N = 302)

| **Food environment domain** | **Mean ± SD** | **Minimum** | **Maximum** | **Interpretation** |
| --- | --- | --- | --- | --- |
| Food Availability (FA) | 2.76 ± 0.52 | 1.40 | 4.20 | Moderate |
| Timing (TA) | 2.82 ± 0.50 | 1.36 | 3.86 | Moderate |
| Affordability (AF) | 2.65 ± 0.51 | 1.27 | 4.00 | Moderate |
| Physical Accessibility (PA) | 2.89 ± 0.50 | 1.40 | 4.80 | Moderate |
| Promotion/Visibility (PV) | 3.27 ± 0.52 | 1.44 | 4.33 | Moderate (highest mean) |
| Food Policy Environment (PE) | 2.99 ± 0.71 | 1.20 | 4.60 | Moderate |

### Assessment of the university food environment

The assessment of the university food environment across the six domains in table 4 showed that students generally perceived the food environment as moderate. Food availability was rated as poor by 64(21.5%) of respondents, while temporal food accessibility was rated as moderate by 240(79.5%) of respondents, with 51(16.9%) reporting poor temporal accessibility. Regarding food affordability, nearly three-quarters 206(74.9%) of respondents perceived affordability as moderate, whereas 64(23.3%) considered it poor and only 5(1.8%) considered healthy foods to be affordable. Physical accessibility also, showed the highest proportion of respondents reporting a moderate environment 248(84.6%), while 42(14.3%) perceived poor accessibility. Food promotion and visibility demonstrated a comparatively more favourable pattern, with 222(78.2%) of respondents reporting a moderate promotional environment and 54(19.0%) reporting a good promotional environment; only 8(2.8%) perceived the promotional environment as poor. For the food policy environment, two-thirds 190(66.0%) of respondents reported a moderate policy environment, whereas equal proportions 49(17.0% each) perceived it as either poor or good.

**Table 4.** Categories of the university food environment among undergraduate students (N = 302)

| **Food environment domain** | **Poor n (%)** | **Moderate n (%)** | **Good n (%)** |
| --- | --- | --- | --- |
| Food availability | 64 (21.5) | 228 (76.8) | 5 (1.7) |
| Temporal food accessibility | 51 (16.9) | 240 (79.5) | 11 (3.6) |
| Food affordability | 64 (23.3) | 206 (74.9) | 5 (1.8) |
| Physical accessibility | 42 (14.3) | 248 (84.6) | 3 (1.0) |
| Food promotion environment | 8 (2.8) | 222 (78.2) | 54 (19.0) |
| Food policy environment | 49 (17.0) | 190 (66.0) | 49 (17.0) |

*Percentages are based on valid responses.*

### Overall Circadian Eating Pattern Score of Respondents

The overall mean circadian eating pattern (CEP) score among the undergraduate students as indicated in table 5, was 2.77 ± 0.54 (range: 1.38–4.38), indicating a moderate circadian eating pattern. The relatively narrow standard deviation suggests moderate variability in meal timing behaviours among respondents. Overall, the findings indicate that while students exhibited some regularity in their daily eating schedules, irregular meal timing and late eating behaviours were still present within the study population. Because the questionnaire uses a 5-point Likert scale, a mean score of 2.77 lies within the moderate category. This suggests that students neither demonstrated consistently healthy circadian eating behaviours nor extremely poor eating timing practices. Instead, the average student reported a mixture of desirable behaviours (such as regular meal consumption) and less desirable behaviours (such as delayed meals, meal skipping, or late-night eating)

**Table 5.** Overall Circadian Eating Pattern Score of Respondents.

**5. Overall Circadian Eating Pattern Score of Respondents**
| Variable | N | Mean ± SD | Minimum | Maximum | Range |
| --- | --- | --- | --- | --- | --- |
| Circadian Eating Pattern (CEP) Score | 302 | 2.77 ± 0.54 | 1.38 | 4.38 | 3.00 |

### The circadian eating patterns of undergraduate students (timing and frequency of meals across the day and night)

The overall mean circadian eating pattern score among the respondents was 2.77 ± 0.54, with scores ranging from 1.38 to 4.38, indicating a moderate overall circadian eating pattern. When categorized, in Fig. 1, 217(71.9%) of the respondents had a moderate circadian eating pattern, 64(21.2%) had a poor circadian eating pattern, and only 21(7.0%) demonstrated a good circadian eating pattern. Overall, these findings indicate that while most undergraduate students exhibited moderately healthy meal timing behaviours, optimal circadian eating practices were uncommon, highlighting opportunities for interventions aimed at promoting healthier meal timing and regular eating schedules. These findings suggest that although most students maintained moderately regular meal timing and eating behaviours, only a small proportion practiced optimal circadian eating patterns, whereas approximately one in five students exhibited poor meal timing behaviours that may have adverse implications for nutritional health.

**Fig. 1:**
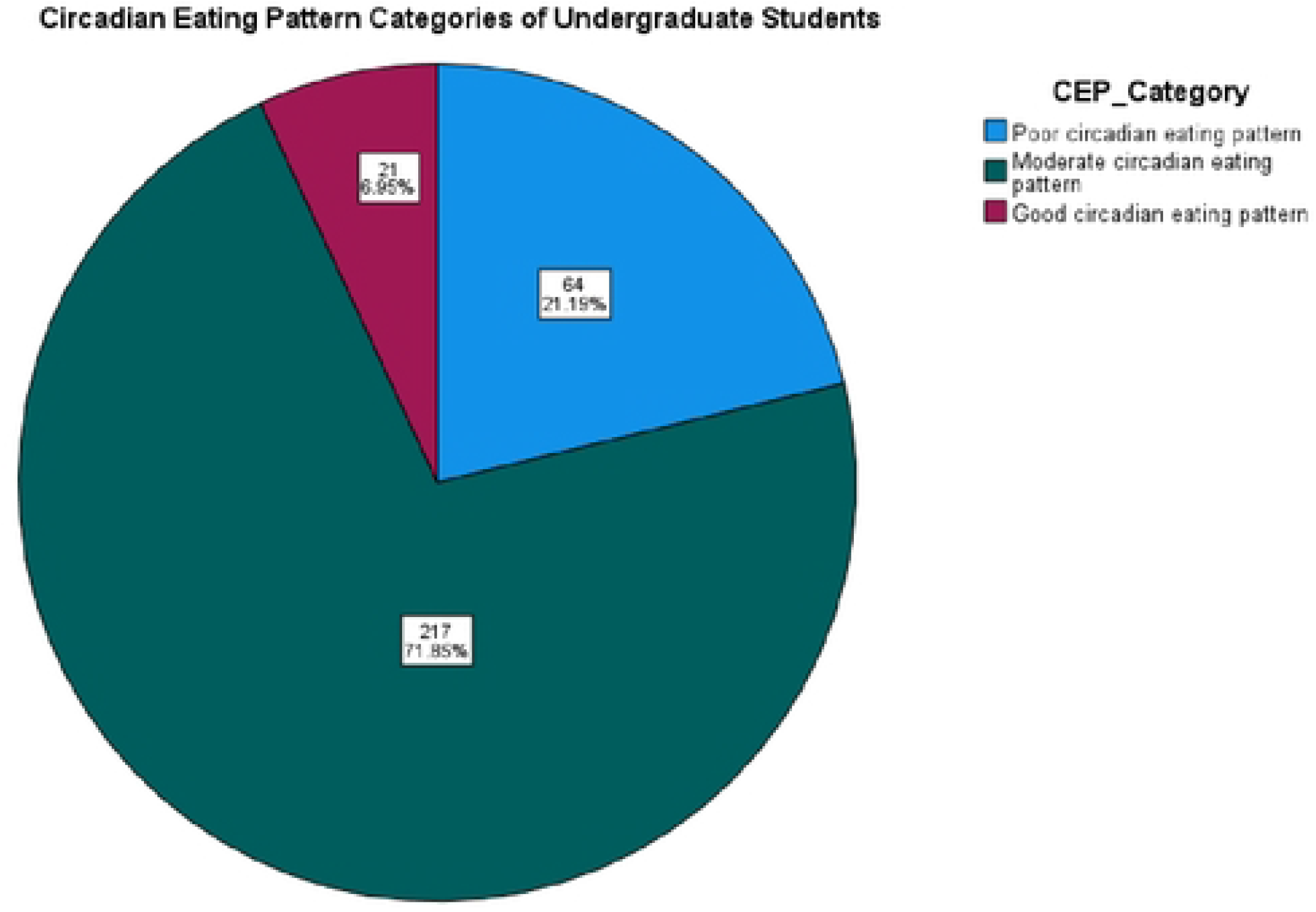
Circadian Eating Pattern Categories of Undergraduate Students

### Nutritional status of undergraduate students

The nutritional status of the respondents was assessed using Body Mass Index (BMI). The findings showed that the majority of the undergraduate students had a normal BMI 254(84.1%). Approximately one in ten students were underweight 28(9.3%), while a smaller proportion were overweight 20(6.6%).

**Fig. 2:**
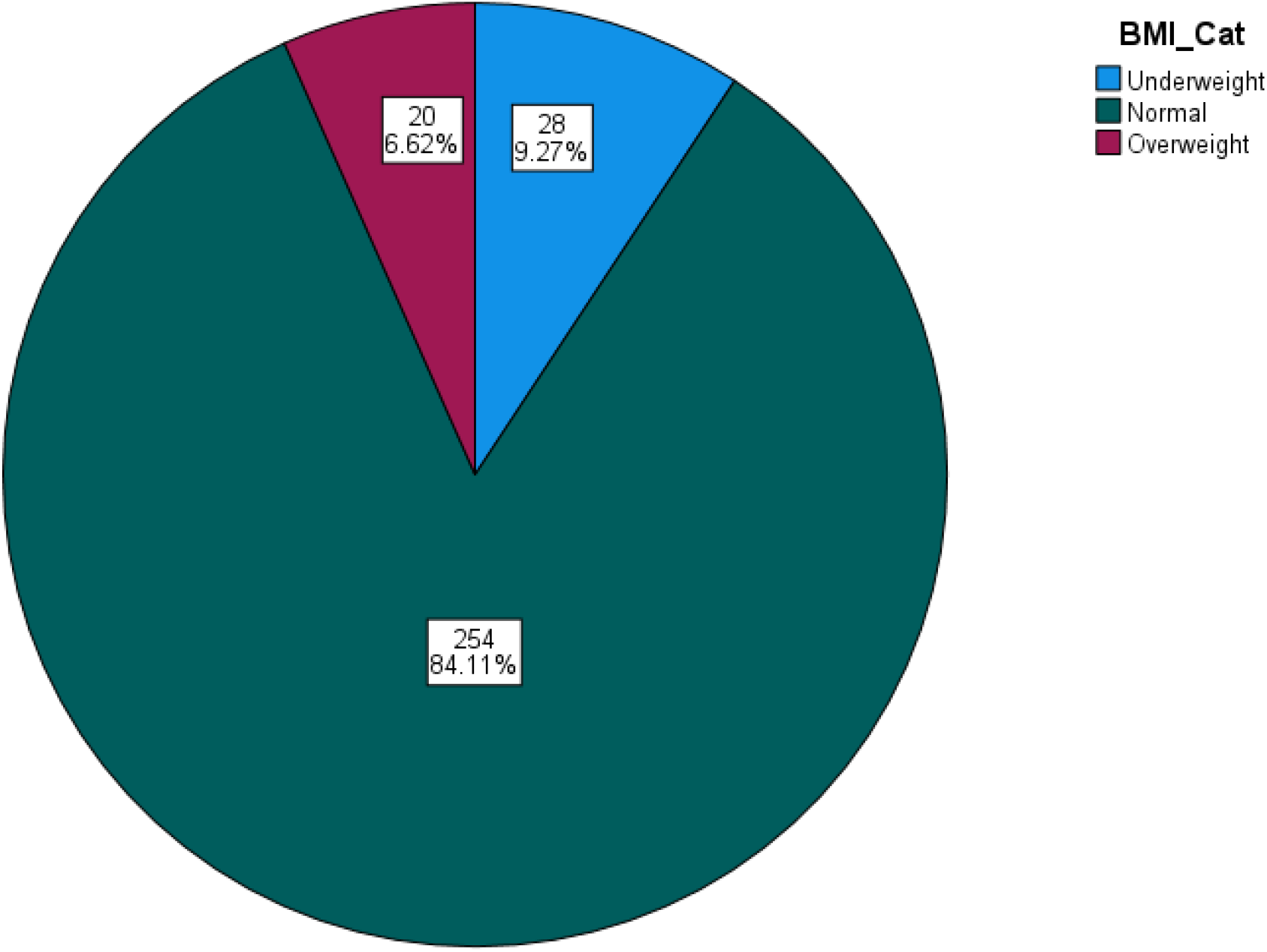
The Nutritional Status of Undergraduate Students

### Factors within the university environment influencing late-night eating or irregular meal timing

The factors within the university environment influencing late-night eating and irregular meal timing are presented in Table 6. The highest mean scores were observed for “*Lack of time makes me depend on fast foods”* (3.80 ± 1.05), “*Academic workload affecting meal timing”* (3.71 ± 1.03), “*the lack of refrigerated storage in hostels prevents from keeping fresh vegetables/fruits for more than a day”* (3.61 ± 1.20), and “*eating at irregular times because they are often held back by academic activities”* (3.59 ± 1.21), indicating that these factors were the most commonly reported influences on students’ eating behaviours. Conversely, “*the academic workload affecting meal timing”* (3.03 ± 1.13) had the lowest mean score, followed by *“skip meals to save money for handouts and academic data”* (3.17 ± 1.17) and *“consuming high-sugar energy drinks because they are the most advertised products on campus billboards/kiosks”* (3.31 ± 1.25).

**Table 6.** Factors within the university environment influencing late-night eating or irregular meal timing (N = 302)

| Variable | Mean ± SD | Interpretation |
| --- | --- | --- |
| My lecture timetable leaves no room for a standard lunch break. | 3.44 ± 1.15 | Moderate influence |
| High academic stress (tests/assignments) triggers my night eating. | 3.50 ± 1.16 | Moderate influence |
| The high cost of healthy food makes me settle for cheap night snacks. | 3.55 ± 1.15 | Moderate influence |
| I skip meals to save money for handouts and academic data. | 3.17 ± 1.17 | Moderate influence |
| The lack of refrigerated storage in my hostel prevents me from keeping fresh vegetables/fruits for more than a day. | 3.61 ± 1.20 | High influence |
| I consume high-sugar energy drinks because they are the most advertised products on campus billboards/kiosks. | 3.31 ± 1.25 | Moderate influence |
| Academic workload affects my meal timing. | 3.71 ± 1.03 | High influence |
| Peer influence affects my eating habits. | 3.03 ± 1.13 | Moderate influence |
| Lack of time makes me depend on fast foods. | 3.80 ± 1.05 | High influence |
| I eat at irregular times because I am often held back by academic activities | 3.59 ± 1.21 | High influence |

Overall, 163(54.0%) of respondents reported a moderate influence of university environmental factors on their eating patterns, while 128(42.4%) reported a high influence. Only 11(3.6%) reported a low influence. These findings suggest that university environmental factors play a considerable role in shaping students’ meal timing and late-night eating behaviours

**Fig 3:**
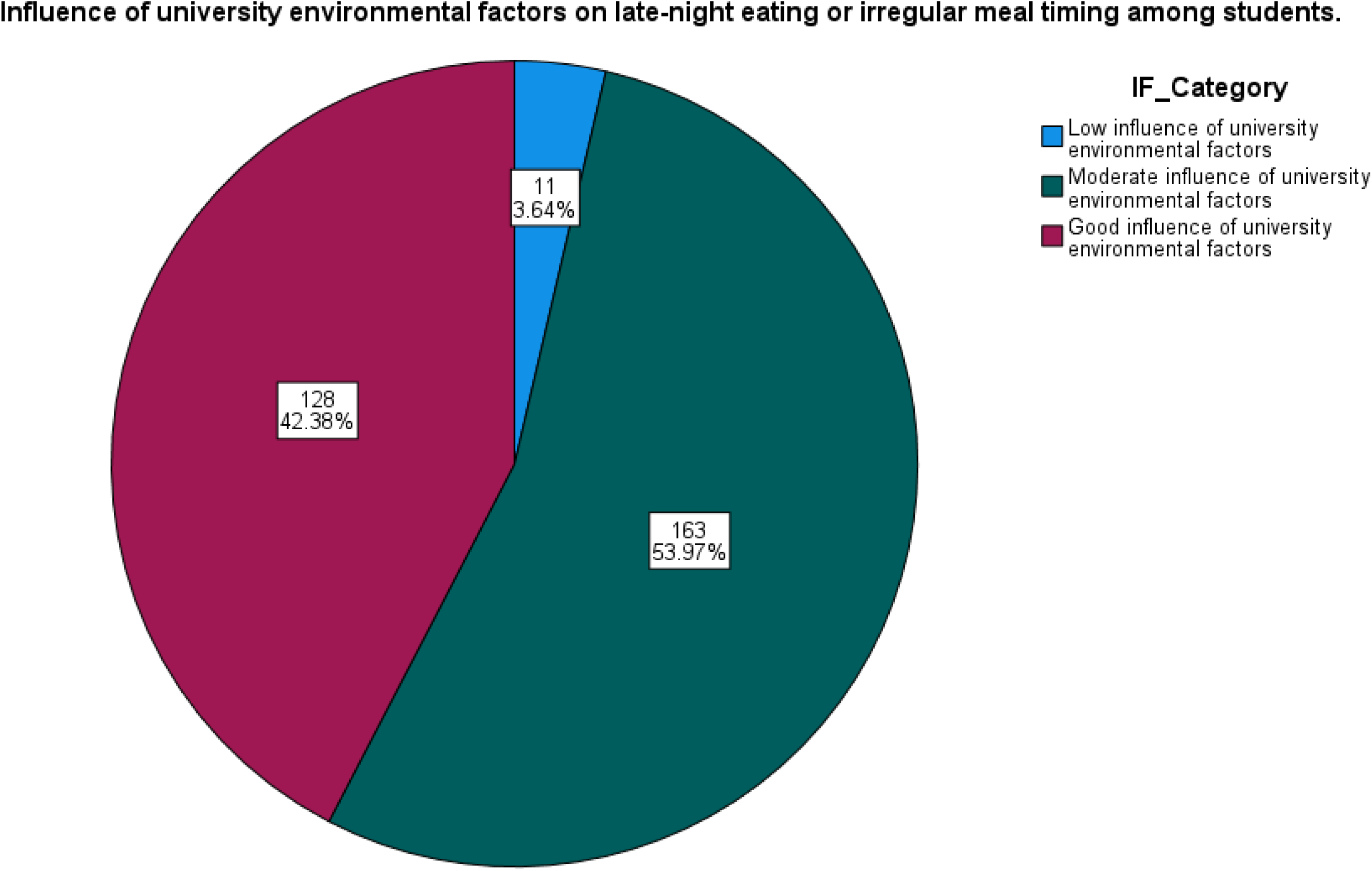
Influence of university environmental factors on late-night eating or irregular meal timing among students.

### Association between Food Environment Domains, Circadian Eating Pattern and Nutritional Status (BMI Category) among Undergraduate Students

Table 7 presents the association between circadian eating pattern, food environment domains and nutritional status among undergraduate students. A statistically significant association was observed between circadian eating pattern and BMI category (χ² = 7.346, *p* = 0.025). Students with poor circadian eating patterns had a higher prevalence of underweight 9(14.1%) and overweight 8(12.5%) compared with those with moderate/good circadian eating patterns, among whom 19(8.0%) were underweight and 12(5.0%) were overweight. Conversely, normal BMI was more prevalent among students with moderate/good circadian eating patterns 207(87.0%) than among those with poor circadian eating patterns 47(73.4%). Among the six food environment domains, physical food accessibility was the only domain significantly associated with nutritional status (χ² = 7.520, *p* = 0.023). Students reporting poor physical food accessibility had a lower prevalence of normal BMI 47(73.8%) and a higher prevalence of overweight 7(16.7%) than those reporting moderate/good physical food accessibility, among whom 214(85.3%) had normal BMI and 13(5.2%) were overweight.

**Table 7.**
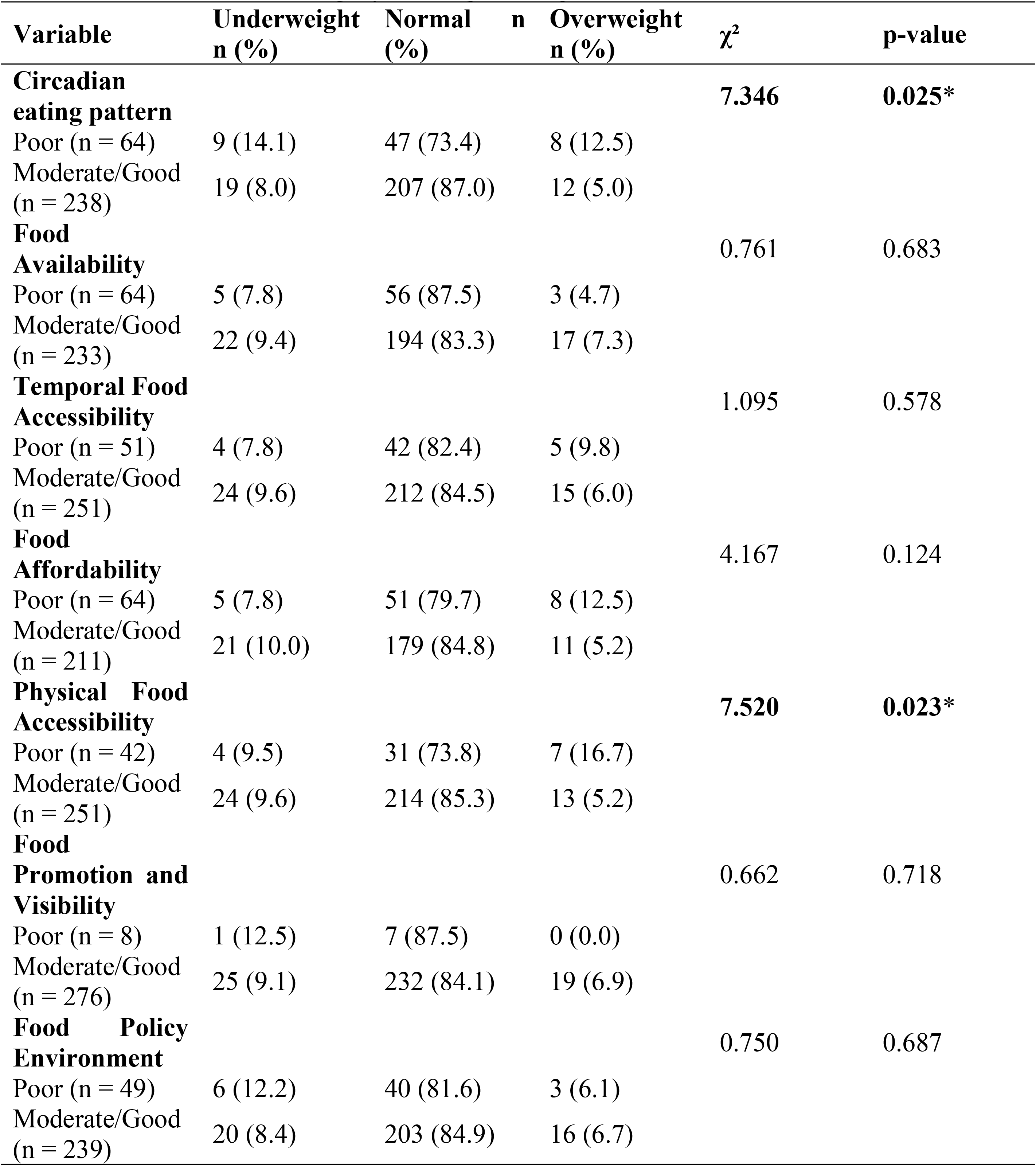
Association between Food Environment Domains, Circadian Eating Pattern and Nutritional Status (BMI Category) among Undergraduate Students (N = 302)

No statistically significant associations were observed between BMI category and food availability (χ² = 0.761, *p* = 0.683), temporal food accessibility (χ² = 1.095, *p* = 0.578), food affordability (χ² = 4.167, *p* = 0.124), food promotion and visibility (χ² = 0.662, *p* = 0.718), or food policy environment (χ² = 0.750, *p* = 0.687).

### Association between University Food Environment and Circadian Eating Patterns of Undergraduate Students

Table 8 presents the association between the university food environment and circadian eating patterns among undergraduate students. A significant association was observed between food availability and circadian eating patterns (χ² = 9.001, *p* = 0.003). Students reporting poor food availability were approximately twice as likely to exhibit poor circadian eating patterns 22(34.4%) compared with those reporting moderate/good food availability 40(17.2%). Temporal food accessibility was also significantly associated with circadian eating patterns (χ² = 28.452, *p* < 0.001). Nearly half 25(49.0%) of students with poor temporal food accessibility had poor circadian eating patterns, compared with only 39(15.5%) among those with moderate/good temporal accessibility.

**Table 8.** Association between University Food Environment and Circadian Eating Patterns of Undergraduate Students (N = 302)

| **Food environment variable** | **Poor CEP n (%)** | **Moderate/Good CEP n (%)** | **$\chi^2$** | **p-value** | **Interpretation** |
| --- | --- | --- | --- | --- | --- |
| **Food Availability** | 22 (34.4) | 42 (65.6) | 9.001 | **0.003** | Significant |
| Moderate/Good availability | 40 (17.2) | 193 (82.8) |  |  |  |
| **Temporal Accessibility** | 25 (49.0) | 26 (51.0) | 28.452 | **<0.001** | Significant |
| Moderate/Good accessibility | 39 (15.5) | 212 (84.5) |  |  |  |
| **Food Affordability** | 28 (43.8) | 36 (56.3) | 29.406 | **<0.001** | Significant |
| Moderate/Good affordability | 27 (12.8) | 184 (87.2) |  |  |  |
| **Physical Accessibility** | 30 (71.4) | 12 (28.6) | 70.611 | **<0.001** | Significant |
| Moderate/Good accessibility | 34 (13.5) | 217 (86.5) |  |  |  |
| **Promotion & Visibility** | 0 (0.0) | 8 (100.0) | 2.395 | 0.122 | Not significant |
| Moderate/Good promotion | 64 (23.2) | 212 (76.8) |  |  |  |
| **Food Policy Environment** | 29 (59.2) | 20 (40.8) | 52.654 | **<0.001** | Significant |
| Moderate/Good policy | 31 (13.0) | 208 (87.0) |  |  |  |
| **Overall Food Environment** | 21 (67.7) | 10 (32.3) | 44.369 | **<0.001** | Significant |

Similarly, food affordability demonstrated a statistically significant association with circadian eating patterns (χ² = 29.406, *p* < 0.001). Poor circadian eating patterns were reported by 28(43.8%) of students experiencing poor food affordability, compared with 27(12.8%) among students with moderate or good food affordability. Physical food accessibility exhibited the strongest association with circadian eating patterns (χ² = 70.611, *p* < 0.001). More than seven out of every ten students 30(71.4%) with poor physical food accessibility had poor circadian eating patterns, whereas only 34(13.5%) of those with moderate/good physical accessibility exhibited poor circadian eating patterns. The food policy environment was also significantly associated with circadian eating patterns (χ² = 52.654, *p* < 0.001). Students reporting a poor food policy environment were substantially more likely to have poor circadian eating patterns 29(59.2%) than those reporting a moderate/good food policy environment 31(13.0%).

In contrast, food promotion and visibility was not significantly associated with circadian eating patterns (χ² = 2.395, *p* = 0.122), indicating that promotional activities and visibility of foods on campus did not significantly influence students’ meal timing behaviors in this study.

Overall, the university food environment showed a strong and statistically significant association with circadian eating patterns (χ² = 44.369, *p* < 0.001). Approximately two-thirds (67.7%) of students exposed to a poor overall food environment exhibited poor circadian eating patterns compared with only 16.0% of those exposed to a moderate or good food environment.

### Univariable and multivariable binary logistic regression analysis of factors associated with poor circadian eating patterns among undergraduate students

Binary logistic regression was performed in Table 9 to identify factors associated with poor circadian eating patterns among undergraduate students. In the univariable analysis, poor food availability (COR = 3.02, 95% CI: 1.59–5.72, *p* = 0.001), poor temporal food accessibility (COR = 6.31, 95% CI: 3.20–12.48, *p* < 0.001), poor food affordability (COR = 5.31, 95% CI: 2.78–10.14, *p* < 0.001), poor physical food accessibility (COR = 14.41, 95% CI: 6.73–30.88, *p* < 0.001), and a poor food policy environment (COR = 8.75, 95% CI: 4.41–17.35, *p* < 0.001) were each significantly associated with increased odds of poor circadian eating patterns. Age, residence, and food allowance were not significantly associated with circadian eating patterns in the univariable analyses (*p* > 0.05). The promotion and visibility domain could not be reliably estimated because of sparse data.

**Table 9.**
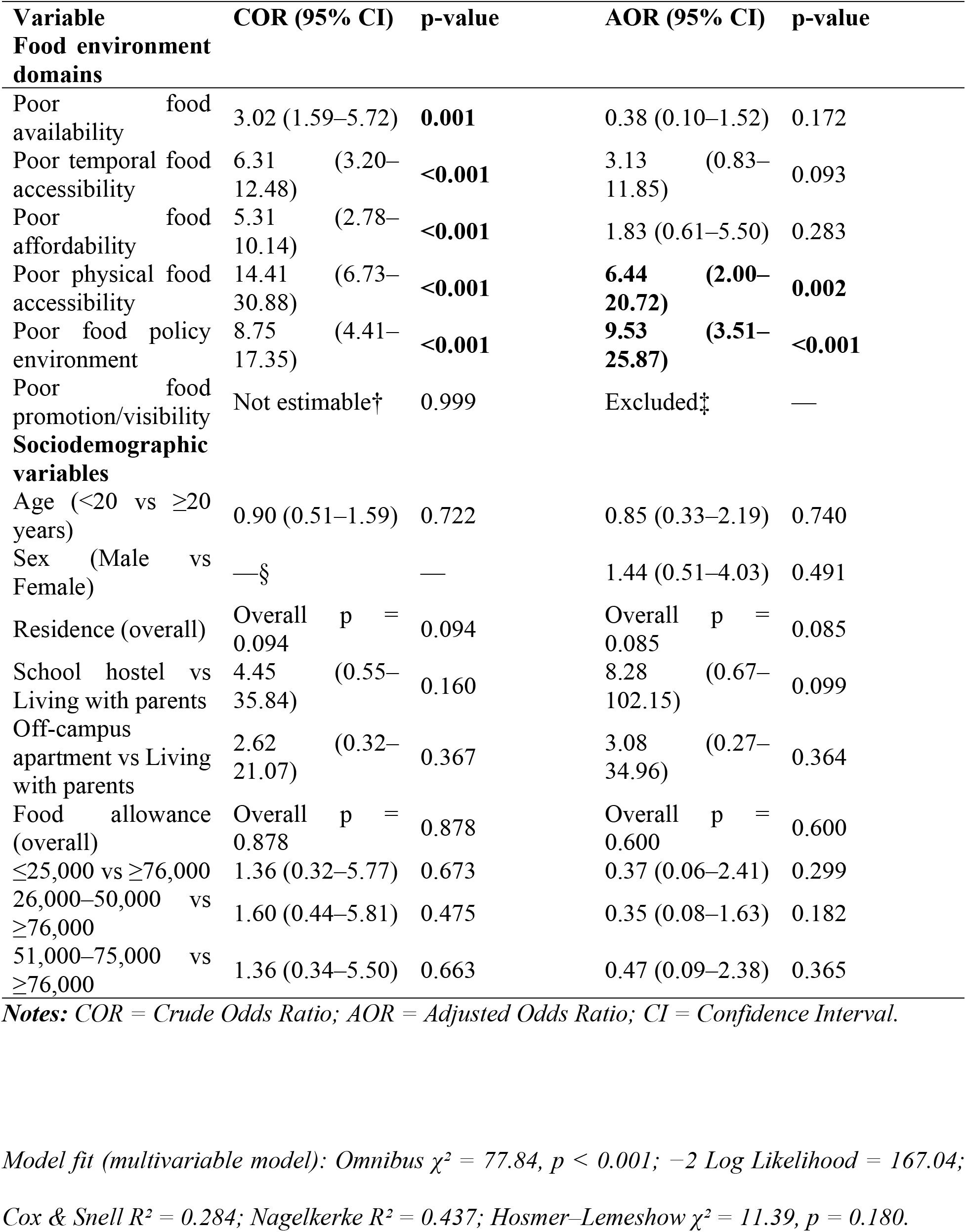
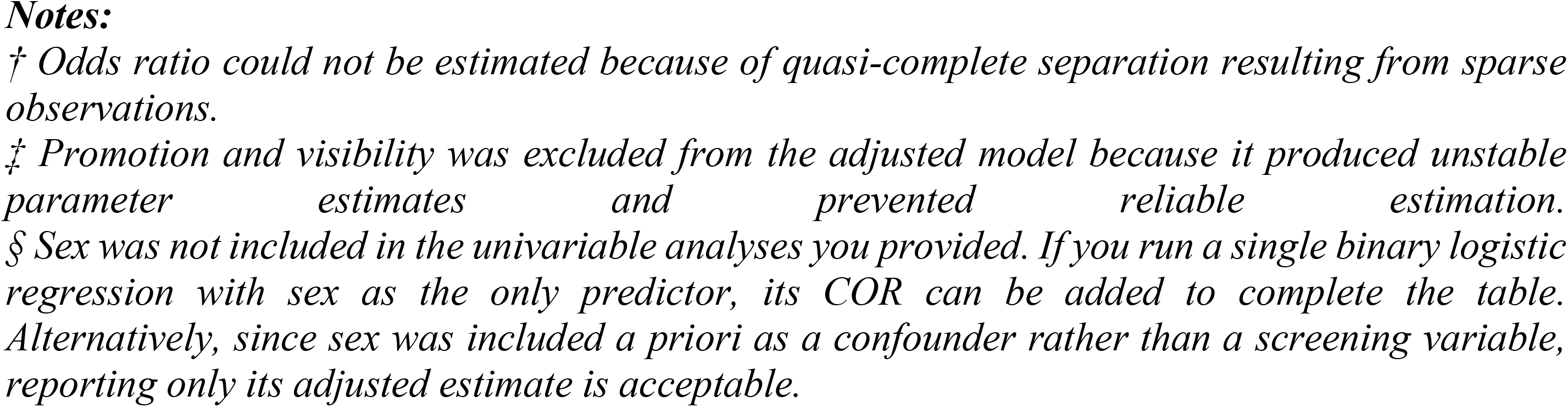
Univariable and multivariable binary logistic regression analysis of factors associated with poor circadian eating patterns among undergraduate students (N = 302)

After adjusting for age, sex, residence, and monthly food allowance, only physical food accessibility and food policy environment remained statistically significant independent predictors of poor circadian eating patterns. Students exposed to poor physical food accessibility had 6.44 times higher odds of exhibiting poor circadian eating patterns than those with moderate/good physical food accessibility (AOR = 6.44, 95% CI: 2.00–20.72, *p* = 0.002). Similarly, respondents exposed to a poor food policy environment had 9.53 times higher odds of poor circadian eating patterns than those experiencing a moderate/good food policy environment (AOR = 9.53, 95% CI: 3.51–25.87, *p* < 0.001). Although food availability, temporal food accessibility, and food affordability were significant in the univariable analyses, these associations were attenuated after adjustment for potential confounders. Age, sex, residence, and monthly food allowance were also not independently associated with poor circadian eating patterns.

### Multivariable binary logistic regression showing factors associated with poor circadian eating patterns among undergraduate students

In table 10, a multivariable binary logistic regression analysis was performed to identify independent predictors of poor circadian eating patterns among undergraduate students. The overall model was statistically significant (Omnibus χ² = 77.84, df = 12, p < 0.001), indicating that the included variables collectively predicted poor circadian eating patterns. The model demonstrated an acceptable fit to the data (Hosmer–Lemeshow χ² = 11.39, df = 8, p = 0.180) and explained approximately 28.4% (Cox & Snell R²) to 43.7% (Nagelkerke R²) of the variation in circadian eating patterns.

**Table 10.** Multivariable binary logistic regression showing factors associated with poor circadian eating patterns among undergraduate students (N = 302)

| Variable | AOR (Exp(B)) | 95% CI | p-value |
| --- | --- | --- | --- |
| Food availability (Poor vs Moderate/Good) | 0.38 | 0.10–1.52 | 0.172 |
| Temporal accessibility (Poor vs Moderate/Good) | 3.13 | 0.83–11.85 | 0.093 |
| Food affordability (Poor vs Moderate/Good) | 1.83 | 0.61–5.50 | 0.283 |
| Physical food accessibility (Poor vs Moderate/Good) | **6.44** | **2.00–20.72** | **0.002** |
| Food policy environment (Poor vs Moderate/Good) | **9.53** | **3.51–25.87** | **<0.001** |
| Age (<20 vs ≥20 years) | 0.85 | 0.33–2.19 | 0.740 |
| Sex (Male vs Female) | 1.44 | 0.51–4.03 | 0.491 |
| Residence (overall) | — | — | 0.085 |
| Food allowance (overall) | — | — | 0.600 |

**Notes:** COR = Crude Odds Ratio; AOR = Adjusted Odds Ratio; CI = Confidence Interval. Model fit (multivariable model): Omnibus $\chi^2 = 77.84$ , $p < 0.001$ ; $-2 \text{ Log Likelihood} = 167.04$ ; Cox & Snell $R^2 = 0.284$ ; Nagelkerke $R^2 = 0.437$ ; Hosmer–Lemeshow $\chi^2 = 11.39$ , $p = 0.180$ .
† Odds ratio could not be reliably estimated because of sparse data (quasi-complete separation).
‡ Promotion and visibility was excluded from the multivariable model because it produced unstable parameter estimates.
§ Sex was included in the adjusted model as an a priori confounder; the univariable estimate was not reported.

After adjusting for age, sex, residence and monthly food allowance, poor physical food accessibility was independently associated with poor circadian eating patterns. Students reporting poor physical food accessibility had 6.44 times higher odds of poor circadian eating patterns compared with those reporting moderate/good physical food accessibility (AOR = 6.44, 95% CI: 2.00–20.72, p = 0.002).

Similarly, respondents exposed to a poor food policy environment were 9.53 times more likely to exhibit poor circadian eating patterns than those exposed to a moderate/good food policy environment (AOR = 9.53, 95% CI: 3.51–25.87, p < 0.001).

Although poor temporal food accessibility was associated with increased odds of poor circadian eating patterns (AOR = 3.13, 95% CI: 0.83–11.85), the association did not reach statistical significance (p = 0.093). Food availability, food affordability, age, sex, residence and monthly food allowance were also not significantly associated with poor circadian eating patterns after adjustment (all p > 0.05).

### Discussion of Findings

This study assessed the university food environment among undergraduate students of the University of Medical Sciences, Ondo. The findings showed that the majority of the respondents had a moderate perception of the university food environment, indicating that although food was generally available and accessible on campus, there were still concerns regarding affordability, availability of healthy food options, operating hours of food outlets, and the overall quality of the campus food environment. These findings suggest that the university food environment provides students with access to food but may not consistently support healthy eating behaviours. The findings are consistent with the report of Almoraie et al. (2025), a narrative review conducted among university students in Jeddah, Saudi Arabia, where it was observed that university students are highly influenced by their food environment, particularly the availability and accessibility of food outlets on campus (27). According to Almoraie et al. (2025), limited availability of healthy food options and easy access to energy-dense, nutrient-poor foods may encourage unhealthy food choices among university students. Similarly, Deliens et al. (2014) reported that university campuses often expose students to food environments that promote convenience foods rather than nutritious meals, thereby increasing the likelihood of poor dietary practices (28). The findings also agree with Turner et al. (2018), who conducted a study in low- and middle-income countries and reported that food availability, accessibility, affordability, and food promotion are major environmental determinants of students’ dietary behaviour (29). Students are more likely to consume foods that are inexpensive, readily available, and easily accessible, regardless of their nutritional quality. This may explain why many respondents in this study reported moderate rather than favourable perceptions of the university food environment. Furthermore, the findings support the ecological model of health behaviour, which explains that food choices are influenced not only by individual preferences but also by environmental factors (30). Since undergraduate students spend a considerable amount of time within the university environment, the availability of healthy foods, pricing strategies, campus food policies, and accessibility of food outlets may significantly influence their eating behaviour. Therefore, improving the university food environment may encourage healthier dietary choices and reduce unhealthy eating practices among students.

The findings of this study further revealed that the majority of respondents demonstrated moderate circadian eating patterns. This indicates that while many students maintained relatively regular meal timing, a substantial proportion still engaged in behaviours such as skipping meals, delaying meals because of academic activities, eating late at night, and consuming food at irregular hours. These behaviours suggest that many undergraduate students do not consistently align their eating patterns with their biological circadian rhythm. This finding agrees with the work of Verde et al. (2024), who reported that university students commonly skip breakfast, consume meals late at night, and have irregular meal schedules due to academic demands and lifestyle factors (31). The authors explained that these behaviours may disrupt the body’s circadian rhythm and increase the risk of metabolic disorders. Similarly, Vujović et al. (2022) found that university schedules often compel students to eat at biologically inappropriate times, resulting in circadian misalignment and poor metabolic health (32). The finding is also consistent with Malin (2021), who conducted a study on the meal Impact on circadian-related health, where it was reported that irregular meal timing, meal skipping, and late-night eating negatively affect glucose metabolism, insulin sensitivity, and overall metabolic function (33). Likewise, Zhang et al. (2025) observed that eating patterns aligned with the body’s biological clock promote better metabolic health, whereas irregular eating patterns increase the risk of overweight, obesity, and chronic diseases (34). The moderate circadian eating patterns observed in this study may be attributed to the demanding academic schedules of undergraduate students, prolonged lecture hours, financial constraints, and limited availability of healthy food options during the day, and easy access to food during late-night study periods. These factors may encourage students to postpone meals, skip breakfast, or consume meals late at night, thereby disrupting normal circadian eating patterns. Generally, the findings indicate that although many students attempt to maintain regular eating habits, university-related environmental and academic factors continue to influence meal timing and contribute to circadian eating disruptions among undergraduate students.

The findings also showed that the majority of respondents had a normal Body Mass Index (BMI), while a smaller proportion were underweight, overweight, or obese. This finding suggests that although most undergraduate students maintained a healthy body weight, a considerable number were either overweight or obese, indicating the presence of nutritional and lifestyle challenges among the study population. The occurrence of overweight and obesity among some respondents may be associated with unhealthy dietary practices, irregular meal timing, physical inactivity, and exposure to an unhealthy university food environment (34). This finding is consistent with the reports of Deliens et al. (2021), who observed that although many university students maintain a healthy BMI, a substantial proportion experience overweight and obesity due to unhealthy eating behaviours developed during university life (35). Similarly, Herforth et al. (2015) reported that inadequate consumption of nutrient-rich foods and increased intake of energy-dense, nutrient-poor foods contribute to weight gain among young adults (36). The finding also agrees with Malin (2021), who reported that irregular eating patterns, particularly meal skipping and late-night eating, may increase the risk of obesity by disrupting glucose metabolism and energy balance (33). Furthermore, Vujović et al. (2022) explained that late isocaloric eating can affect hunger, energy expenditure, and metabolic pathways. The predominance of respondents with normal BMI may be attributed to the relatively young age of undergraduate students and their higher levels of daily physical activity, including walking between lecture halls and participating in academic activities. However, the presence of overweight and obesity among some respondents suggests that interventions promoting healthy eating behaviours and appropriate meal timing are still necessary to prevent future increases in obesity among university students.

The findings further revealed that academic workload, lack of time, poor accessibility to healthy foods, late operating hours of food vendors, and the availability of food during late-night study periods were the major factors influencing late-night eating and irregular meal timing among undergraduate students. These findings indicate that both academic and environmental factors significantly influence students’ eating behaviours. This finding is consistent with Vujović et al. (2022), who reported that late eating can be influenced by factors related to daily schedules and may have implications for circadian alignment and metabolic health (18). Similarly, Deliens et al. (2021) reported that university food environments characterised by the easy availability of convenience foods and limited healthy alternatives may encourage unhealthy eating habits. Turner et al. (2021) further explained that food accessibility and affordability are important factors influencing food choices. Students may postpone meals because of lectures and later rely on readily available fast foods or snacks during evening and nighttime hours (10). The findings also support the work of Boyland et al. (2022), who found that exposure to food marketing and the widespread availability of unhealthy foods can encourage the consumption of energy-dense foods (37). The influence of peers, academic stress, and limited time for meal preparation may further reinforce unhealthy eating behaviours among university students. The findings imply that improving the university food environment through increased availability of healthy foods, appropriate regulation of food outlet operating hours, nutrition education, and implementation of supportive food policies may help reduce irregular eating patterns and encourage healthier meal timing among undergraduate students (37).

The findings of this study showed a statistically significant association between the university food environment and circadian eating patterns. This implies that characteristics of the university food environment, particularly food accessibility and food policy, significantly influence the timing and regularity of meals among undergraduate students. This finding supports previous studies which reported that campus food environments play an important role in determining students’ dietary behaviours and meal timing (38). The findings of this study also indicated that aspects of the university food environment were associated with respondents’ nutritional status. This finding suggests that the availability, accessibility, affordability, and quality of foods within the university environment may influence students’ body weight over time by affecting their dietary choices and eating behaviours. These findings agree with previous reports that unhealthy food environments contribute to overweight and obesity among university students by encouraging the consumption of energy-dense foods and discouraging healthier dietary practices (15).

### 5.2 Conclusion

This study assessed the influence of the university food environment on the circadian eating patterns of undergraduate students at the University of Medical Sciences, Ondo. The findings revealed that most respondents perceived the university food environment as moderate, indicating that while food was generally available and accessible, there were concerns regarding the affordability, accessibility, and availability of healthy food options. The study further established that the majority of the respondents exhibited moderate circadian eating patterns, although irregular meal timing, meal skipping, and late-night eating were common among a considerable proportion of the students. Academic workload, limited time for meal preparation, easy access to food during late-night study periods, and poor accessibility to healthy food options were identified as the major factors influencing irregular eating behaviours.

Most respondents had a normal Body Mass Index (BMI), although the presence of overweight and obesity among some students suggests that unhealthy dietary practices remain a public health concern. Furthermore, the study demonstrated that the university food environment significantly influences students’ circadian eating patterns and nutritional status. In particular, food accessibility and campus food policies were identified as important determinants of healthy meal timing and eating behaviours. Overall, the study concludes that improving the university food environment through better availability and accessibility of healthy foods, supportive food policies, and nutrition education will promote healthier circadian eating patterns and contribute to improved nutritional status among undergraduate students.

### 5.3 Recommendations

Based on the findings of this study, the management of the University of Medical Sciences, Ondo, should develop and implement comprehensive campus food policies that promote the availability, affordability, and accessibility of healthy food options within the university environment. University food vendors should be encouraged to provide nutritious and balanced meals, including fruits, vegetables, whole grains, and other healthy food choices at affordable prices. Regular nutrition education programmes and health promotion campaigns should be organised to educate students on healthy eating, regular meal timing, the importance of aligning eating patterns with the body’s circadian rhythm, and the need to minimise late-night eating and meal skipping. Healthy food outlets should be strategically located within the university to improve students’ physical access to nutritious foods, while the operating hours of food outlets should be reviewed to ensure that healthy meal options are available throughout the day. Students should also be encouraged to plan their meals despite academic commitments and adopt healthy lifestyle practices, including regular physical activity and adequate sleep. Finally, future researchers should conduct similar studies in other universities using longitudinal study designs to provide stronger evidence on the long-term influence of university food environments on circadian eating patterns and obesity.

## Data Availability

Data is available upon request from the corresponding author and already provided as part of the submitted article.

## Abbreviations

CEP: Circadian Eating Pattern
BMI: Body Mass Index
WHO: World Health Organisation

## Consent for publication

Not applicable.

## Availability of data and materials

Data is available upon request from the corresponding author.

## Authors’ Contributions

Oluwabunmi Emmanuel Folorunso, Joy Chinaza Amadi, Oluwaseun Funm Akinmoladun and Beatrice Oguba all made substantial contributions to the conceptualisation and design of the study. Each author participated actively in drafting the manuscript and provided critical intellectual input during the revision process. All authors approved the submission of the manuscript to the current journal, granted final approval of the version designated for publication, and agreed to be accountable for all aspects of the work.

## Acknowledgements

The authors wish to express sincere gratitude to all individuals whose support contributed to the successful completion of this study, particularly the participants who gave their time and the research assistants who provided invaluable assistance throughout the data collection process.

